# Formative usability assessment of a new connector design for peritoneal dialysis

**DOI:** 10.1101/2022.03.31.22273212

**Authors:** Ibrahim Yekinni, Tom Viker, Bill Hartsig, Arthur Erdman

**Affiliations:** Bakken Medical Devices Center, University of Minnesota, Minnesota, USA; Cerovations LLC, Minnesota, USA; Primordial Soup LLC, Minnesota, USA

## Abstract

**Background:** This paper presents a first, formative study to explore the usability of a new peritoneal dialysis connector design intended for use by patients. The study was conducted with a user population of both naive users and experienced peritoneal dialysis patients across a range of ages. The goals of the study were to evaluate the usability of the key user interfaces of this connector design by test participants representative of new and experienced peritoneal dialysis patients, as well as to evaluate the use of the connector as it interacts with other components of the peritoneal dialysis system including peritoneal dialysis fluid bags and tubing. Further objectives were to capture any usability issues and obtain participants’ feedback on the design.

**Methods:** A total of 7 patient and non-patient participants received brief training and performed simulated connection and disconnection of peritoneal catheter extension sets for therapy with the new design.

**Results:** All 7 participants completed the simulated connection and disconnection tasks successfully, with only one use error (0.22%), 18 close calls (4.0%), 6 use difficulties (1.3%) observed from the total of 449 use steps performed by all participants. Other findings include usability improvement with repeated use, participants feedback and suggestions for the ‘protective enclosure’, a novel feature of the touchless connector design.

**Conclusion:** The studied connector design showed minimal use errors or difficulties and based on participant feedback, the usability can be significantly improved with minimal modifications in future prototypes.

## 1. INTRODUCTION

End-stage renal disease affects millions of people around the world and peritoneal dialysis is one of the options available to patients for its treatment. In peritoneal dialysis, dialysis fluid is passed through an implanted catheter into a space within the abdomen called the peritoneum and the membranes of this space serve as a filter through which excess fluid and solutes are removed from the blood.

Peritoneal dialysis has advantages in cost effectiveness compared to the alternative (Hemodialysis) and may help improve access to therapy in low resource settings. The International Society of Nephrology has recommended a Peritoneal Dialysis first approach as a strategy for dealing with the limited access to renal replacement therapy in low and middle income countries where up to 7 million deaths may be occurring due to poor access to therapy [1, 2]. In the United States, a new government initiative, Advancing American Kidney Health Initiative, has set a goal to increase the proportion of kidney failure patients on home treatments like peritoneal dialysis from current ∼10% to about 80% by 2025, with new changes in payment models to support this [3].

However, a major drawback of peritoneal dialysis is peritonitis, an infection of the peritoneal membrane through which the therapy occurs. Peritonitis leads to repeat episodes in 3% - 20% (14% overall), surgeries to remove the peritoneal catheter in 10% - 88% (22% overall), permanent transfer to hemodialysis in 9% - 74% (18% overall) and death in 0.9% - 8.6% (2% - 6% overall) [4-7]. While there are different sources of peritonitis, a significant and preventable cause is contamination of the peritoneal catheter while patients connect for therapy. This could be through touch contamination, which contributes to 40% of peritonitis episodes. Other sources include respiratory droplets when patients connect for therapy without masks, environmental contaminants e.g. from pets, etc [8,9]. Over 50% of the bacteria resulting in peritonitis is from bacteria colonizing the skin or mucous membranes, supporting the importance of touch and respiratory contamination [10]. Current strategies for prevention train patients in aseptic techniques but adherence is poor and over 50% of patients stop prescribed practices within 6 months of commencing therapy [11].

A more effective elimination of touch, aerosol or environmental contamination without overdependence on patient adherence may help reduce peritonitis episodes and this is the theory behind the touchless connector system evaluated in this study. **Figure 1** shows the components of the touchless connector system along with a common peritoneal dialysis system used in standard practice. While most components of the touchless connector system remain the same as in the standard of care, the unique features that confer its advantages mean that it will be used by patients differently. It is therefore important to ensure that interaction between patients and the touchless connector system results in safe, reliable and effective peritoneal dialysis therapy. The use of the touchless connector system should be understandable, intuitive and reproducible with minimization of use-related hazards and risks.

**Figure 1.**
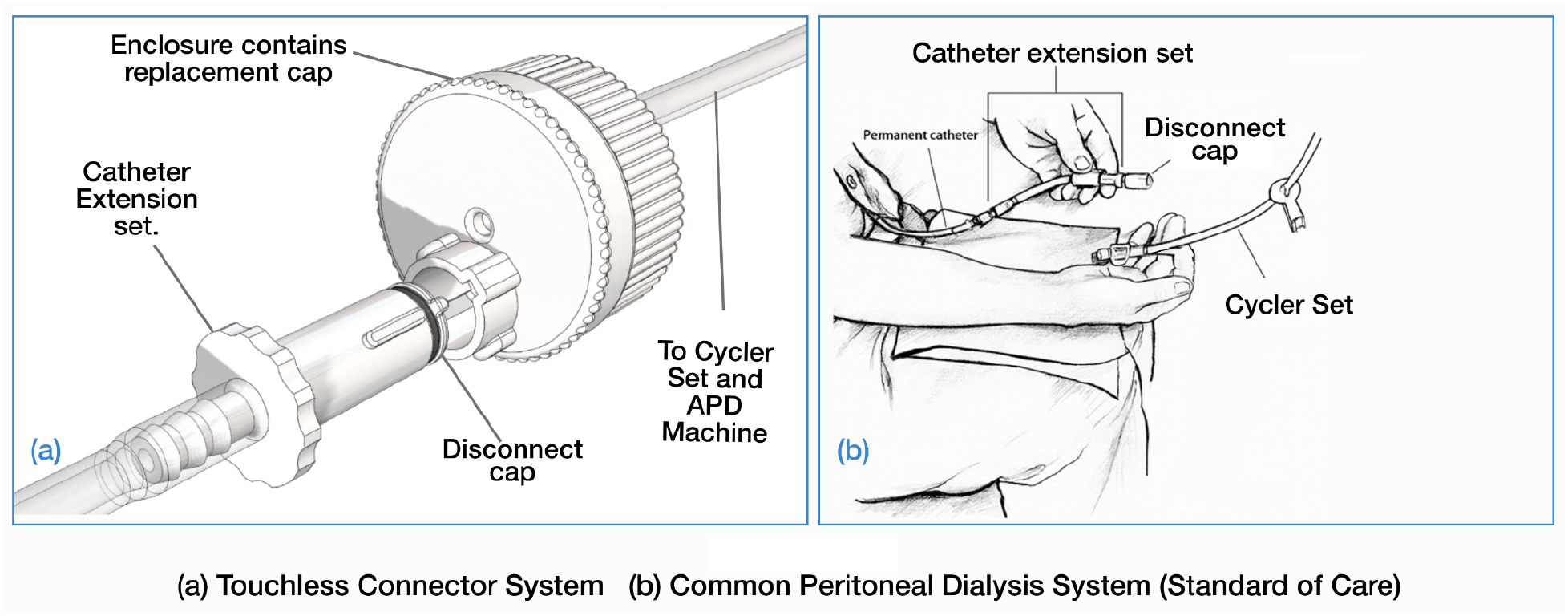

## 2. PROBLEM STATEMENT

In this paper, we describe a formative study evaluating the usability of the touchless connector system, identifying use errors that may require risk mitigation, and the modifications implemented in response to patient-user feedback.

## 3. MATERIALS AND METHODS

### 3.1 Test Articles and Study Materials

Materials for use in this study include:

Prototypes of the touchless connector system;

IV Pole;

Permanent peritoneal catheter simulator;

Tubing sets with clamps and bags;

Video camera to capture a wide view of participant sessions;

Printed copies of the moderator’s workbook and consent/confidentiality forms.

### 3.2 Study Design

The primary objective was to evaluate the usability of the key user interfaces of the touchless connector system by test participants representative of intended users. The secondary objective was to evaluate the use of the touchless connector system as it interacts with the rest of the peritoneal dialysis system (peritoneal dialysis fluid bags and tubing).

Factors evaluated include:

1. Ergonomics, the device size and grip interaction;
2. Ease of use and intuitiveness of the device design through form language and control style;
3. Visual, auditory, and/or tactile feedback communicating successfully completed connections.

Participants were recruited through a commercial service, Fieldwork Minneapolis (Edina, MN) and the Kidney Center of the University of Minnesota Masonic Children’s Hospital. A purposive sampling approach was used to capture a range of demographic characteristics and minimize bias where possible. Human study exemption was granted by the University of Minnesota Institutional Review Board (STUDY00010480) and participants granted their written informed consent.

### 3.3 Study Procedure

The study was conducted on separate days at two locations - University Enterprise Labs (Saint Paul, MN) and the University of Minnesota Masonic Children’s Hospital’s Kidney Center (Minneapolis, MN). BH served as the moderator, administering the usability test sessions and recording the test data. All interview sessions were video recorded and photographed from multiple angles to capture use scenarios of interest i.e., use errors, close calls, etc.

A permanent peritoneal catheter simulator **(Figure 2)**, made from an apron containing an intravenous solution bag connected with a catheter extension set was used. There was also an Intravenous solution bag hanging from an IV pole with the enclosure end of the touchless connector system. Participants used the permanent peritoneal catheter simulator and the enclosure of the touchless connector system to simulate the connection and disconnection procedures performed by patients in peritoneal dialysis therapy.

**Figure 2.**
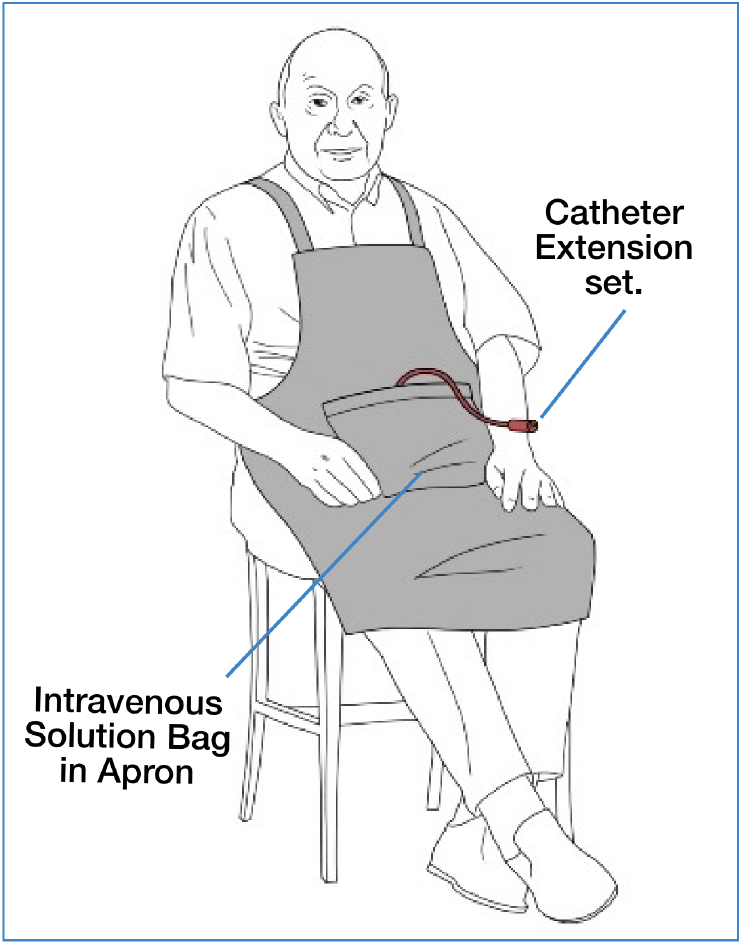
Permanent Peritoneal Catheter Simulator

Each test session was initiated by asking demographic questions, introducing the non-patient participants to peritoneal dialysis and explaining the purpose of the test. Participants were then presented with a prototype of the touchless connector system. TV trained participants with multiple prototypes and after training, they were allowed to repeat connection and disconnection procedures about six times. Initial attempts were with the help of instructions on a whiteboard, and the final attempts were without the instructions. After each task and during a post-task interview, the moderator, BH, interviewed the participants about their interactions with the device. The participants were also asked to provide feedback on specific aspects of their experience using the device.

### 3.4 Use Scenarios Evaluated

Participants were instructed to perform tasks in two use scenarios (1) Connecting permanent peritoneal catheter for therapy; (2) Disconnecting permanent peritoneal catheter after completing therapy. These tasks/scenarios, together with the individual steps required in each, are outlined in **Table 1**.

**Table 1:** Use Scenarios and Individual Steps Evaluated.

| **Table 1: Use Scenarios and Individual Steps Evaluated** |  |
| --- | --- |
| **1. Connecting catheter for therapy** | **2. Disconnecting catheter after therapy** |
| (i) Insert transfer set into enclosure | (i) Close valve to stop flow |
| (ii) Twist the transfer set to unlock cap | (ii) Pull transfer set to disconnect from fluid bag |
| (iii) Pull the transfer set to remove cap | (iii) Rotate enclosure base $\frac{1}{4}$ turn to next detent |
| (iv) Rotate enclosure base $\frac{1}{4}$ turn | (iv) Push transfer set down to add new cap |
| (v) Push transfer set in to connect with fluid bag | (v) Twist transfer set to lock new cap |
| (vi) Open valve to allow flow | (vi) Withdraw transfer set |

In both scenarios, the participants wear the permanent catheter simulator with the catheter extension set extending out of the abdominal area of the apron.

### 3.5 Data Collection and Analysis

Data collection was limited to qualitative data coming from participant comments, moderator observations during sessions and observations during review of video recordings. Focus was on acceptance and understanding of required steps taken, successful task completions, potential usability errors, issues, or frustrations, and additional opinions or comments. The data was then categorized to identify common themes and concept directions that may be preferred by users. The moderator notes and video recording sessions were analyzed to identify use errors that may affect the ability of the touchless connector to perform effectively and safely.

## 4. RESULTS AND DISCUSSION

### 4.1 Quantitative Findings

A total of 7 naive participants who had no previous exposure to the touchless connector system participated in the study. 5 of these were non-dialysis patients recruited from the general public, while two were peritoneal dialysis patients at the Kidney Center of the University of Minnesota Masonic Children’s Hospital. The demographic and other characteristics of participants are summarized in **Table 2**.

**Table 2:** Participant demographics.

| Participant No | Gender | Age | Limitations | Dialysis Patient? |
| --- | --- | --- | --- | --- |
| 01 | Male | 51 - 60 | None | No |
| 02 | Male | 71 - 80 | Visual | No |
| 03 | Male | 21 - 30 | None | No |
| 04 | Female | 21 - 30 | None | No |
| 05 | Female | 61 - 70 | Dexterity | No |
| 06 | Male | 11 - 20 | None | Yes |
| 07 | Female | 11 - 20 | None | Yes |

Only 3 of the 7 enrolled participants completed all steps in the two evaluated use scenarios six times. In the rest of the sessions, test prototypes malfunctioned due to repeated use requiring discontinuation of steps or entire use scenarios. 2 participants had one or more steps discontinued with replacement of the prototype while the remaining two had entire use scenarios discontinued due to worn out prototypes leaving a total of 449 use steps for analysis. Of the 449 analyzed steps, there were 18 (4.0%) self-corrected close calls, 6 (1.3%) use difficulties, and 1 (0.22%) use error. There were no significant indicators that any user interactions posed a threat to safety or efficacy. All key events are summarized in **Figure 3** with explanation of each in the following sections.

**Figure 3:**
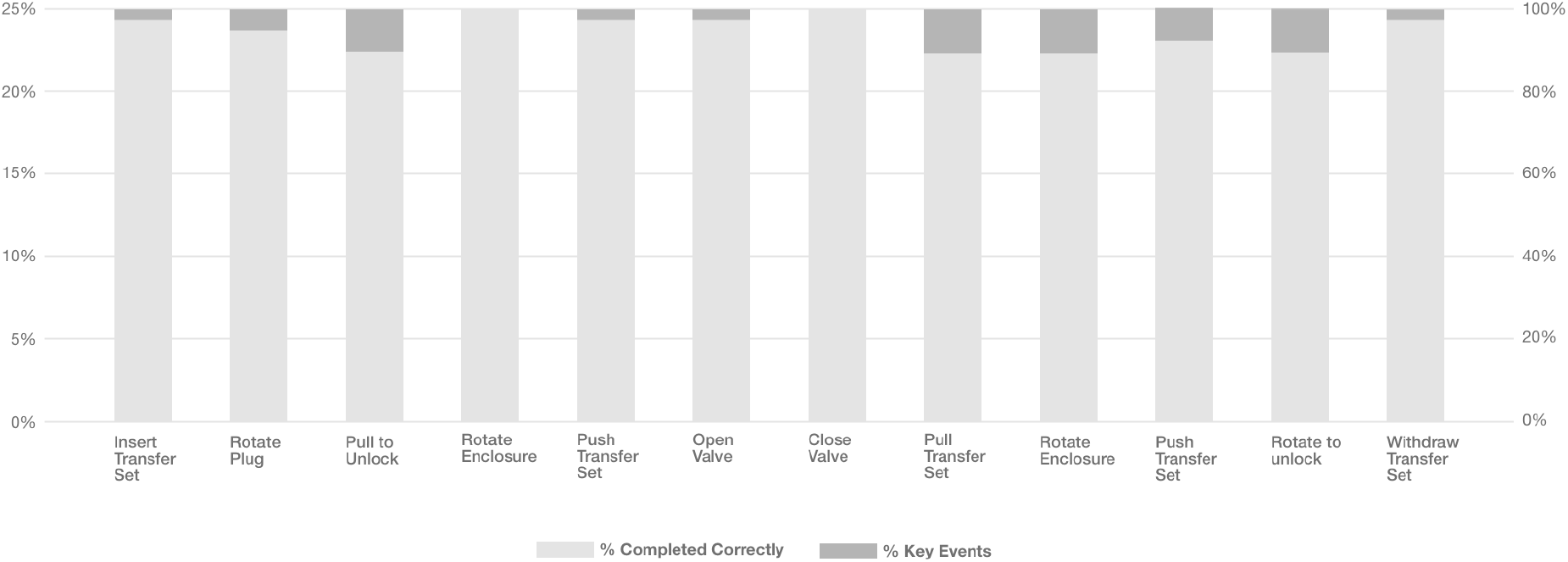
Summary of 449 total steps from all participants

#### 4.1.1 Close calls

These were the cases in which the participant almost commits an error but “catches” himself or herself in time to avoid making the error. It also includes cases where a participant commits a use error but detects it in short order and recovers before the error becomes consequential. In total there were 18 close calls identified in all 7 participants, 4.0% of the 449 total steps. These close calls were distributed across 10 of the 12 use steps being investigated. The only use steps without close calls were (1) *Rotating Enclosure*; and (2) *Closing Valve*. (**Figure 4**)

**Figure 4:**
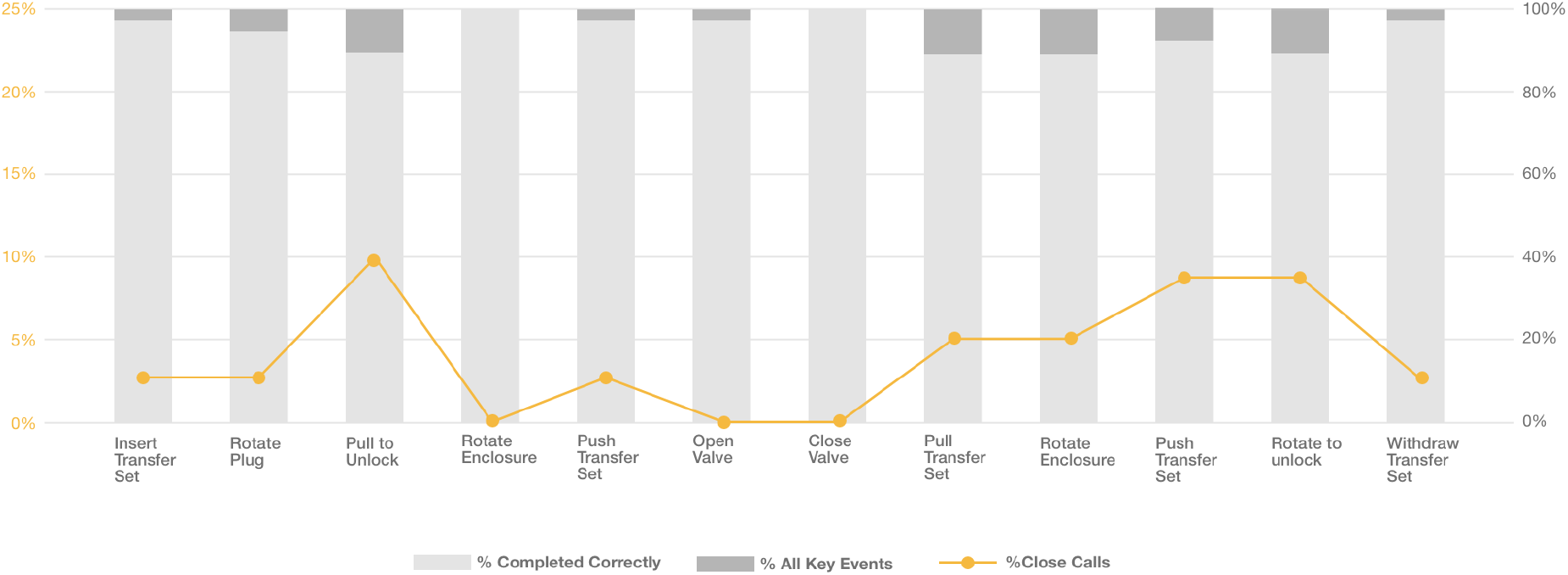
Close calls

#### 4.1.2 Use difficulties

These were the cases in which the participants appeared to struggle to perform a task, indicated by multiple attempts to perform the task, anecdotal comments about the task’s difficulty, requests for assistance with the task, facial expressions suggesting frustration/confusion and longer than usual task performance times.

In total, there were 6 use difficulties identified for 3 of the participants. The use difficulties observed made up 1.3% of the 449 total use steps attempted by the participants. These use difficulties were distributed across 5 of the 12 use steps being evaluated. These steps were (1) *Rotate Plug*; (2) *Open Valve*; (3) *Pull catheter extension set*; (4) *Rotate Enclosure*, during disconnection; (5) *Rotate to unlock*. (**Figure 5**).

**Figure 5:**
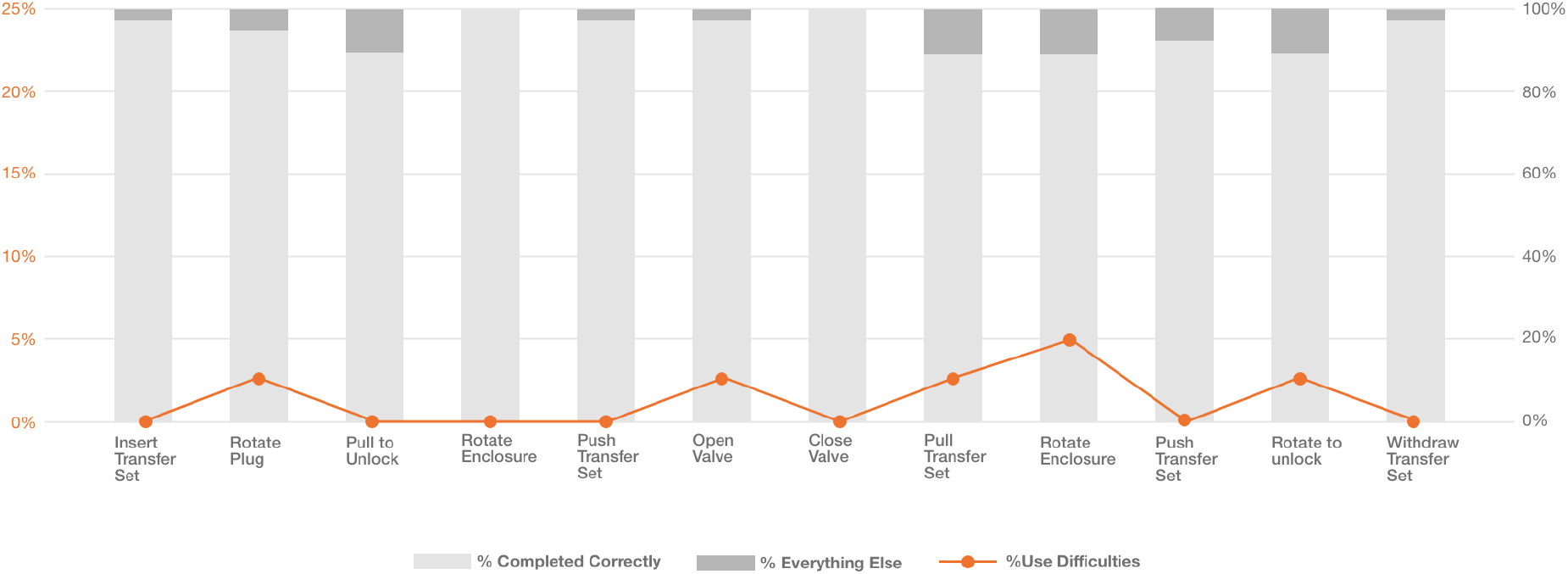
Use difficulties

#### 4.1.3 Use errors

These were cases in which participants performed a task in an incorrect manner that will not lead to the intended outcome. They may result from incorrect action, or failure to act when action is needed. There was only one use error from one participant, 0.2% of the 449 use steps. It occurred in the “*Pull catheter extension set*” step of the disconnection task (**Figure 6**).

**Figure 6:**
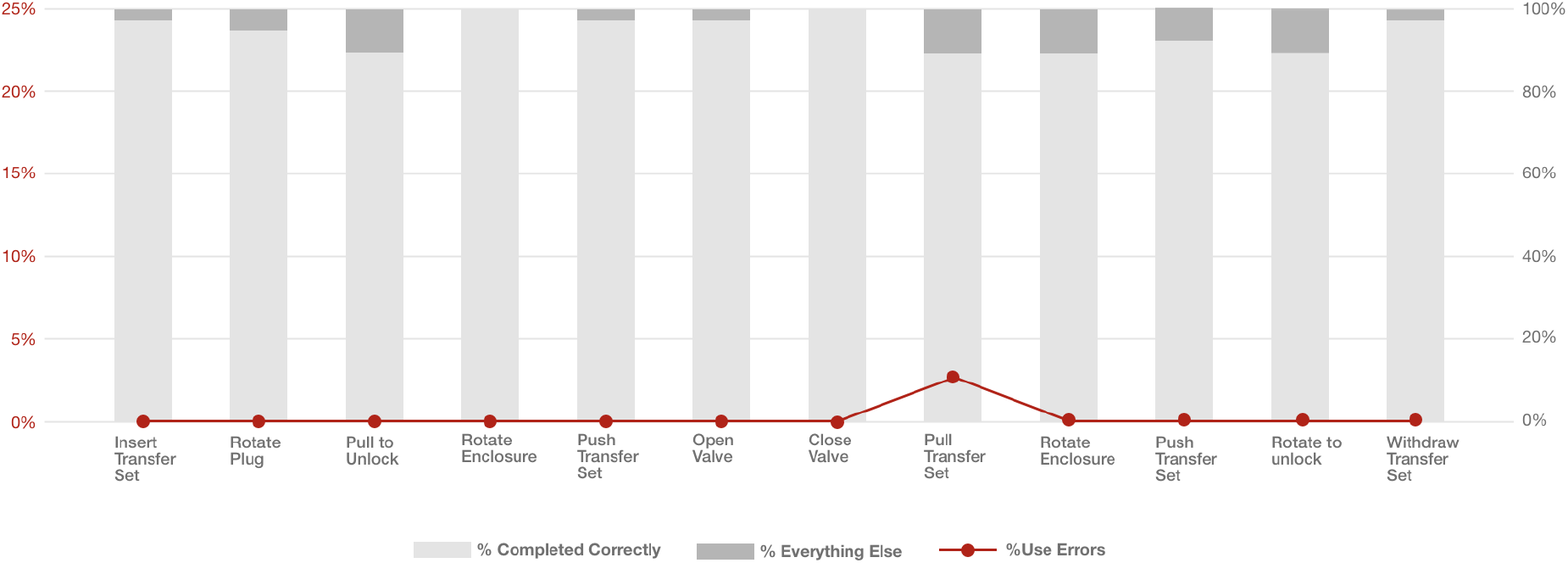
Use errors

### 4.2 Qualitative Findings

BH documented subjective feedback provided by participants and responses to questions asked to understand their thinking while using the device. The following are the recurring themes identified and are described in more detail below.

#### 4.2.1 Usability improves with repeated use

4 of the 7 participants commented on how use improves with subsequent attempts and nearly all participants showed improvement in subsequent attempts. This was demonstrated by them no longer needing the instructions or fewer events (close calls or use difficulties) in subsequent attempts.

*Participant 01* commented, “*It’s easier the second time around*” as he moved through the steps more quickly than in his first attempt and seems sure that it’ll become routine with enough use, as he commented: “*Pretty simple. The more you do it the better you get at it*.”

For *participant 02*, he needed the instructions for the first attempt but didn’t by the sixth attempt and he commented “*If you have to do this daily then you’ll learn it pretty quick*.”

*Participant 03* commented after 2 attempts, “*If I did this a couple more times, I’d have the hang of it*.” In the third attempt, he had a much smoother run through as he became more familiar with the device. In the remaining three attempts, he performed all the steps correctly with no use difficulties. The experience was also similar for *participant 05* who was able to perform steps correctly on the third attempt and felt she had improved with practice during the previous two.

#### 4.2.2 Auditory and tactile feedback

Most participants mentioned how they depended on auditory and tactile feedback while performing “*Rotate Enclosure”* and “*Push catheter extension set”* steps of the connection task. These helped them know, while rotating, if they were in the correct position to push the catheter extension set. They believed it would help if the sound were louder and the tactile feedback was stronger. Some participants rotated past the connection point for the catheter extension set when they couldn’t get tactile or auditory feedback that they were in the middle position. Some of the participants also used the alignment of the catheter extension set and fluid bag tubing to know when they’ve reached the middle point.

*Participant 01* mentioned that the click at the middle position was helpful to indicate proper position. It was faint but he could also feel it. He also felt confident the catheter extension set was in the correct position by using the alignment of the catheter extension set and fluid bag tube as an indicator. He relies on the click and tactile feedback to align the middle position and is concerned a noisy environment might cover up the click. *Participant 02* asked about people who might have hearing impairment. He says the click is very quiet and he also cannot feel any tactile sensation with the click. He prefers a stronger and louder clicking sound. *Participant 03* could hear and feel the click at the center position but doesn’t think he would feel it if he rotated faster. He says having both the clicking sound and feeling it is nice for reassurance since he is confident that the connection is correct when he hears the click.

*Participant 06* used the audible click to know when he was at the center position but did not get any tactile feedback with the click. *Participant 07* over-rotated directly into the final position because there was no click.

#### 4.2.3 Enclosure

Participants made various comments about the enclosure design and changes that may be required, including (1) changing terminology for parts in the instructions to make it easier to understand the instruction; (2) providing visual aids to help identify the rotating components and to know the right direction for rotation, i.e. clockwise or anticlockwise; (3) Improving the grip of the rotating component i.e., the ‘plug’ and ‘cap’.

##### 4.2.3.1 Changing terminology of parts in instructions

Participant 01 suggested including in the instructions that it is the ‘cap’ that should be rotated clockwise, since, “*It would be easy to make the wrong assumption*”. In his initial attempt, he turned the ‘plug’ instead of the ‘cap’ because he thought the cap referred to the catheter extension set and didn’t realize it referred to the slim side of the enclosure. Participant 03 also mentioned how he got confused by the terminology of cap/plug in the instructions. For participant 04, the confusion in plug/cap terminology may have resulted in observed use difficulty during the disconnect task of her first attempt. When she got to the *Rotate Enclosure* step (in instructions as ***Rotate cap 1/4 turn***), she was observed pausing to think of this step, and grabbing the grip on the catheter extension set (in instructions as plug) and trying to rotate it. She attempted other maneuvers until she handed back the device thinking it malfunctioned. After TV indicated which part was plug and cap, she said she was confused about the terminology used to describe the parts in this step. She also suggested labeling the parts of the device. Participant 05 had a similar experience, holding the catheter extension set to rotate instead of the enclosure, resulting in confusion over the next steps. She also suggested changing the instructions. Participants 06 and 07 also both rotated the plug instead of the cap.

##### 4.2.3.2 Visual aids for rotating enclosure component and rotation direction

Participant 01 suggested noting in the instructions that the ‘cap’ rotation is clockwise, since, “*It would be easy to make the wrong assumption*”. He also suggested making the rotating component of the enclosure, the ‘cap’, a different color. “*Sets it apart so you know it’s two separate pieces*”.

Participant 02 said “*One thing that my brain hasn’t figured out yet is that you only turn (the cap) clockwise*”. He wants to turn back during the disconnection task as if he is reversing the process for the connection task. “*Once I’m done. I want to go back*”. He also suggests using a different color for the ‘cap’.

Participant 04 suggested the ‘cap’ could be “*a little thicker, it could have a rim on it*” as she points to the perimeter where the grip is. She also suggested the ‘cap’ be a different color and a marker be included on the side of the ‘cap’ to indicate alignment in tandem with a click sound.

##### 4.2.3.3: Improving the grip of rotating components

Participant 01 thought that the grip all round was fine and the ‘cap’ was easy to turn. Participant 02 thinks the ‘plug’ comfort and ease of use is fine, given he’s not wearing his reading glasses but can still see the parts sufficiently to insert the plug in the hole. He also thinks the grip is fine given he has big hands and can still grip it with comfort. However, he doesn’t like the ‘cap’ (rotating component of the Enclosure). “*The disk with the grip is way too small, it’s hard to see, difficult to grab”*. For participant 03, during the first attempt, he felt that the grip on the plug was too small, “*jagged*” and rough on his hands, but was fine with the grip on the cap. However, by his fifth attempt, he thought the grip texture of the plug was more acceptable as he uses it and as he grips lighter. “*It’s less of an annoyance now*”, he said. Participant 05 who has a dexterity limitation thinks that people with arthritis may have trouble with the plug grip. She has braces for each of her wrists. She was wearing the brace for the left hand when she arrived, but she removed it before the test. She mentioned she does not have arthritis, but a condition similar to carpal tunnel syndrome that is exacerbated by typing. She believed she’ll have trouble gripping the plug if her symptoms were worse.

### 4.3 Mitigations

Based on the patterns of user difficulty and feedback from findings, the design of the touchless connector system and the instructions were updated to mitigate those difficulties or improve suggested features as shown in **Table 4**. Updates include changes to terminologies describing catheter extension set and enclosure components in Instructions for Use as suggested by participants, improving auditory and tactile feedback to help participants know they’ve reached the midpoint and should stop during rotation, and modifying design of rotating components to improve grip strength during rotation. The updated design is shown in **Figure 7**.

**Table 4:** Usability issues identified and mitigations.

| Table 4: Usability issues identified and mitigations |  |  |
| --- | --- | --- |
| S/N | Usability Issue Identified | Mitigations |
| 01 | Issues with Instruction | Changes to terminology for parts |
| 02 | Weak auditory and tactile feedback | The design was modified to increase click sound and vibration for improved feedback. |
| 03 | Enclosure issues: |  |
| 3.1 | Terminology of parts in instructions | Parts terminology will be changed in instructions |
| 3.2 | Visual aids for enclosure components and rotation direction | Integrated directional graphics will be considered in future prototypes to indicate direction of rotation of enclosure |
| 3.3 | Grip of rotating components | Design in future prototypes will be modified to improve grip |

### 4.4 Conclusions

This formative usability study sought to evaluate several features of the Touchless Connector System for potential applications in peritoneal dialysis therapy. Overall, the prototypes were found to be usable, with use error and use difficulties mostly around the “Pull Transfer Set” and “Rotate Enclosure” steps when participants attempted disconnecting after simulated therapy. In conclusion, the studied Touchless Connector System design showed minimal use errors or use difficulties and based on participant feedback, the usability can be significantly improved with minimal modifications in future prototypes.

## Data Availability

All data produced in the present study are available upon reasonable request to the authors

## Contributions

IOY drafted the work and this was then revised critically by all authors. All authors were involved in the assessment with TV training the participants and BH being the moderator. All authors contributed to the conception and design of the work. All authors had final approval of the version to be published and are jointly accountable for all aspects of the work.

